# Changes in population immunity to omicron SARS-CoV-2 variants and selected Sarbecoviruses from 2020 to 2023 in urban Colombo, Sri Lanka

**DOI:** 10.1101/2024.10.03.24314822

**Authors:** Farha Bary, Maneshka Vindesh Karunananda, Chandima Jeewandara, Saubhagya Danasekara, Dinuka Guruge, Rizna Rizan, Inoka Sepali Aberathna, Thushali Ranasinghe, Heshan Kuruppu, Jeewantha Jayamali, Lahiru Perera, Harshani Chathurangika, Amaya Gunaratne, Naduni Dasanthi, Chathura Ranatunga, A.W. Shashini Ishara, Sathsara Yatiwelle, Ruwan Wijayamuni, Tiong Kit Tan, Alain Townsend, Graham S. Ogg, Gathsaurie Neelika Malavige

## Abstract

**Background:** To understand how the population immunity evolved over time and possible susceptibility of the Sri Lankan population to emerging SARS-CoV-2 variants, we proceeded to evaluate the changes in antibody positivity rates to omicron variants BA.2.75 and XBB.1.5 and for selected sarbecoviruses.

**Methods:** The haemagglutination test (HAT) was carried out to determine the presence of antibodies against the RBD of the SARS-CoV-2 omicron variants XBB.1.5 and BA.2.75 and the RBD of the Sabecoviruses RaTG13, WIV1, Khosta-2 and SARS-CoV-1, in individuals aged 5 to 80 years of age in years 2020 (n=381), 2022 (n=432) and 2023 (n=382).

**Results:** The highest positivity rates for BA.2.75, RaTG13, WIV1, Khosta-2 and SARS-CoV-1 were seen in 2022, with positivity rates significantly declining to many of the viruses except XBB.1.5 and Khosta-2 by 2023. The positivity rates for Khosta-2 (p<0.001) and WIVI (p<0.001) were significantly lower in children <14 years age, but not for XBB.1.5, BA.2.75 and RaTG13. Children <14 years who were SARS-CoV-2 unvaccinated had the lowest positivity rates for all tested viruses except BA.2.75. <20% of individuals in all age groups had antibody titres equivalent to 1:80, which correspond to neutralising antibody titres by 2023.

**Conclusions:** Population immunity to omicron SARS-CoV-2 variants and selected sarbecoviruses had significantly declined in Colombo, Sri Lanka by 2023. Therefore, although T cells might still offer some protection against severe disease, immunizing vulnerable individuals in the community with protective vaccine designs, might be important to consider at this stage.

## Introduction

Despite widespread immunization and infection, SARS-CoV-2 continues to evolve and cause outbreaks in many parts of the world [1]. Lineages which are descendants of JN.1 variant, such as KP.3, KP.2 and LB.1 are currently the dominant variants globally [1, 2]. Therefore, monovalent vaccine containing JN.1 lineages have been recommended as the COVID-19 vaccine booster vaccines [3]. Both rolling out of booster doses and natural infection maintain high level of population immunity, thereby preventing severe disease leading to hospitalization and mortalities [4, 5]. SARS-CoV-2 is known to rapidly evolve escaping pre-existing neutralizing antibodies generated by previous infecting variants and vaccines [5, 6]. It was recently shown that the evolution of SARS-CoV-2 was dependent on population immunity [5]. Although booster vaccines have been rolled out in many countries, the Sri Lankan population did not receive any vaccines containing the recent XBB.1.5 or JN.1 variants. Sri Lanka has not deployed any COVID-19 vaccines since mid-2022. Furthermore, although there are currently many antivirals treating vulnerable patients with COVID-19 [7], these are not available in Sri Lanka and many other low-middle income and low-income countries.

The number of reported COVID-19 cases does not reflect the actual number of infections in a community, as many have mild or asymptomatic infection [8, 9]. Furthermore, since active surveillance for SARS-CoV-2 is not carried out in many countries, SARS-CoV-2 infections causing outbreaks may be missed as the clinical features mimic infections with influenza, dengue and other febrile illnesses, which can have a similar clinical presentation [10]. Therefore, in order to understand population immunity to SARS-CoV-2 and its emerging variants and other potential sarbecoviruses, it would be important to evaluate the seropositivity of antibodies against these viruses in the population.

The Colombo Municipality Council has a high population density (20,187.8 individuals/km^2^), and experienced high rates of infections throughout the COVID-19 pandemic [9, 11]. We had documented early infection with SARS-CoV-2 in 2020 in the community[12] and subsequently obtained samples in years 2022 and 2023 from this area, to determine changes in population immunity over time. In this study, we proceeded to evaluate the changes in antibody positivity rates to omicron variants BA.2.75 and XBB.1.5 and for selected sarbecoviruses to understand how the population immunity evolved over time and possible susceptibility of the Sri Lankan population to emerging SARS-CoV-2 variants. We further sought to investigate if comorbidities such as diabetes and obesity, influenced the positivity rates for these variants.

## Methodology

### Participants and ethics approval

Assays were carried out on archived samples of individuals aged 5 to 80 years, obtained in the years 2020 (n=381), 2022 (n=432) and 2023 (n=382), following informed written consent. The cohorts in 2020 were recruited 2 months following the initial detection of SARS-CoV-2 in Sri Lanka. Adult individuals recruited in years 2022 and 2023 had received two or more COVID-19 vaccines, while children <14 years had not received the vaccines.

Ethics approval for these studies was obtained by the Ethics Review Committee of the University of Sri Jayewardenepura. The blood samples were obtained from these individuals during these time points for analysis for the presence of SARS-CoV-2 specific antibodies and antibodies to arboviruses. The assays were carried out on samples which had been stored in - 80°C.

Demographic details and information regarding the presence of comorbidities (presence of diabetes, hypertension and chronic kidney disease) were obtained using an interviewer administered questionnaire at the time of recruitment to the study. Anthropometric measures were obtained from individuals recruited in the year 2023. The weight of individuals was obtained using a digital scale with an accuracy of ±0.1kg, while wearing light clothing and being barefooted. The height was measured using a stadiometer with an accuracy of ±0.1cm, and the waist circumference was measured using a non-stretchable tape between the lowest rib and the iliac crest after an exhalation. Females with a waist circumference of ≥80cm and males with a waist circumference of ≥90cm were classified has having central obesity[13]. Those with a BMI below 18.5 kg/m^2^ were classified as underweight, 18.5-23.8 kg/m^2^ normal weight, 23.9-27.5 kg/m^2^as overweight and above 27.5 kg/m^2^ as obese [14].

### Haemagglutination test (HAT) to detect antibodies to the receptor binding domain (RBD) of omicron and selected Sarbecoviruses

The HAT assay was carried out as previously described [15] to determine the presence of antibodies against the RBD of the SARS-CoV-2 omicron variants XBB.1.5 and BA.2.75 and the RBD of the Sabecoviruses RaTG13, WIV1, Khosta-2 and SARS-CoV-1. The assays were carried out and interpreted as previously described at serum dilutions of 1:40 and 1:80 [16, 17]. A known positive sample with 1:80 HAT titre was used as the positive control. A titre of 1:40 was considered as a positive response, while a titre of 1:80 an indicator for the presence of antibody titres which can neutralize the tested SARS-CoV-2 variant as previously described [18]. A HAT titre of 1: 80 was shown to detect 99% of samples which had neutralizing antibody titres of ≥ 20 (50% inhibitory concentrations, IC_50_) when compared to the gold standard microneutralization assay, for the original SARS-CoV-2 virus [18].

### Statistical analysis

GraphPad Prism version 10.1 was used for statistical analysis. As the data were not normally distributed, differences in means were compared using the Mann-Whitney U test (two tailed), and the Freidmans test was used to compare the differences of the antibody levels between different age groups, for different omicron variants and Sarbecoviruses. A post hoc analysis for the relationship between the BMI and a high waist circumference was carried out using the for chi square test setting the standardized residual value at 0.05 significance level, with the alpha level adjusted by using Bonferroni correction.

## Results

### Population immunity to Omicron variants and selected Sarbecoviruses from 2020 to 2023

The positivity rates (HAT titre of 1:40) and presence of HAT titres equivalent to the original neutralising antibody titres (HAT titre of 1:80), were measured in the cohorts in 2020, 2022 and 2023 in different age groups. The number of individuals included in each age group, in each year is shown in supplementary table 1.

In 2020, the positivity rates for omicron variants XBB.1.5 was <10.53% in all age groups and for BA.2.75 <8.2%. While the positivity rates for Khosta-2 and WIVI were <12% in all age groups, the positivity rates for SARS-CoV-1 and RatG13 were higher (Figure 1A). Interestingly, the positivity rates for the omicron variants and the Sarbecoviruses were significantly different between the different age groups (p=0.01), with the highest positivity rates, especially for SARS-CoV-1 and RaTG13 being observed in those >66 years of age.

**Figure 1.**
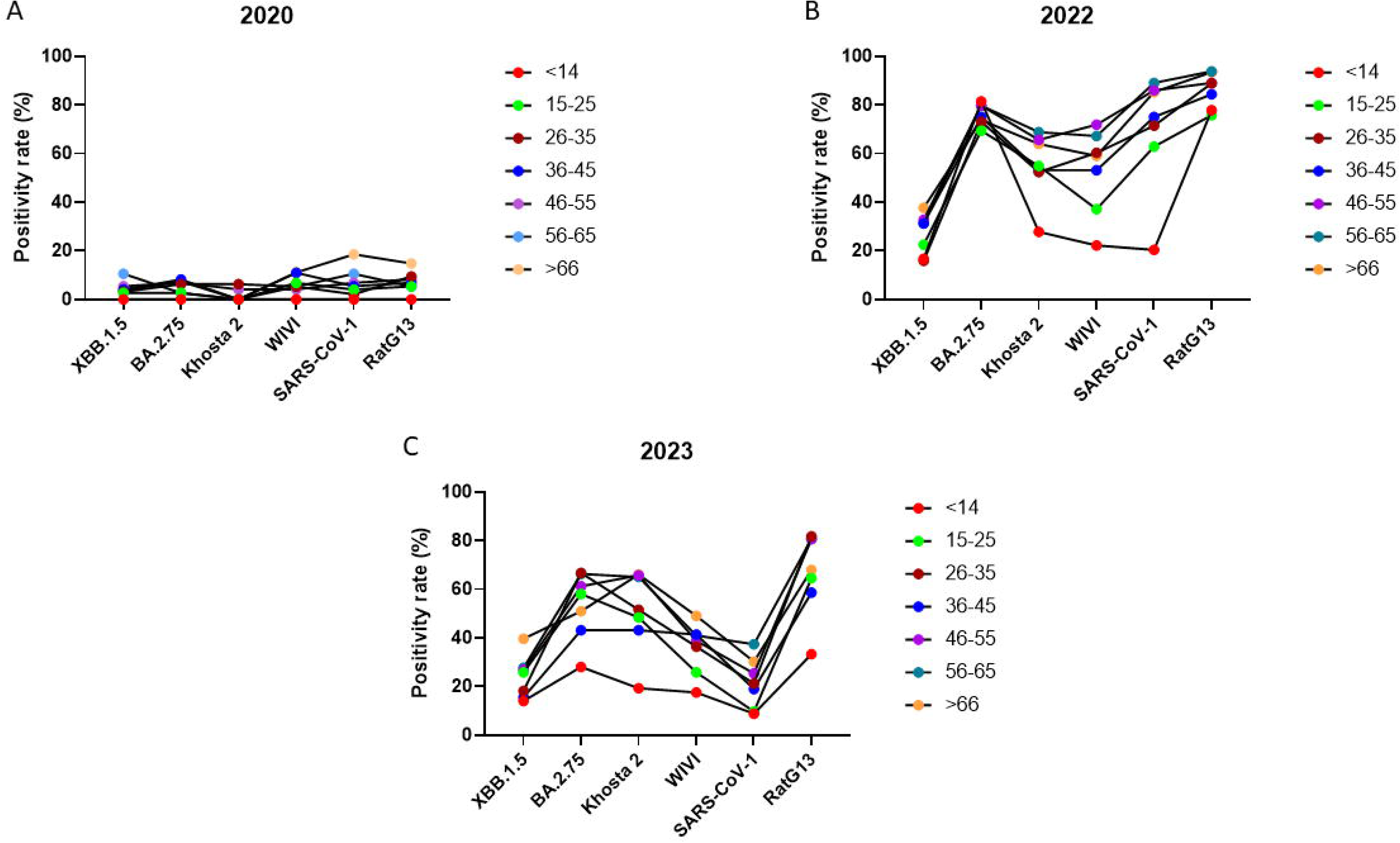
Positivity rates of individuals to two omicron variants and four sarbecoviruses in years 2020, 2022 and 2023. Antibody titres were measured against the receptor binding domain (RBD) of XBB.1.5, BA.2.75, Khosta-2, WIV1, SARS-CoV-1 and RaTG13 in 2020 (n=381), 2022 (n=432) and 2023 (n=382) using the HAT assay in different age groups in individuals aged 5 to 80 years of age (supplementary table 1). Freidman’s test was used to compare changes in the antibody levels between the different age groups, for each omicron variant or sarbecovirus, in 2020 (A), in 2022 (B) and in 2023 (C).

In 2022, the positivity rates for omicron variants and the Sarbecoviruses significantly rose in all age groups, following natural infection and vaccination of the population. However, the positivity rates for Khosta-2 (p<0.001) and WIVI (p<0.001) were significantly lower in children <14 years age, but not for XBB.1.5, BA.2.75 and RaTG13 (Figure 1B). The highest positivity rates were observed for RaTG13 in all age groups, with positivity rates >75%, with individuals aged >55 years reporting positivity rates >93%. The positivity rates were significantly lower (p<0.001) for the omicron variant XBB.1.5, in all age groups compared to RatG13.

In 2023, again the highest positivity rates were seen for RatG13, with positivity rates ranging from 33.3% in children <14 years to 80.6% in older individuals (Figure 1C). By 2023, children <14 years had the lowest positivity rates to both omicron variants and the Sarbecoviruses. The positivity rates were lowest for SARS-CoV-1. The highest positivity rates for RatG13 and BA.2.75 was seen in the 26 to 35 years age group, while the highest positivity rates for Khosta-2 was seen in the >46 years age groups (Figure 1C).

### The changes in the positivity rates for the omicron variants and selected Sarbecoviruses over time

The main SARS-CoV-2 variant that circulated towards the end of 2022 in Sri Lanka was BA.2.75, while this was gradually replaced by XBB.1.5 and the descendants of this lineage in year 2023[2]. The individuals >20 years of age also were offered the BNT162b2 (Pfizer) as the booster dose, which was taken up by approximately 37% of the population by June 2022 [19].

Therefore, as expected the highest positivity rates were seen for BA.2.75 and the sarbecoviruses of clade 1A (RaTG13) clade 1B (W1VI and SARS-CoV-2) and clade 3 (Khosta-2) in all age groups (Figure 2). Interestingly, except for children <14 years of age, in 2022, individuals of other age groups had equal positivity rates for clade 1A sarbecoviruses, which consist of SARS-CoV-2 and its variants and RatG13 along with viruses of clade 1B, and clade 3, and thereby suggesting a broadly cross-reactive immune response.

**Figure 2.**
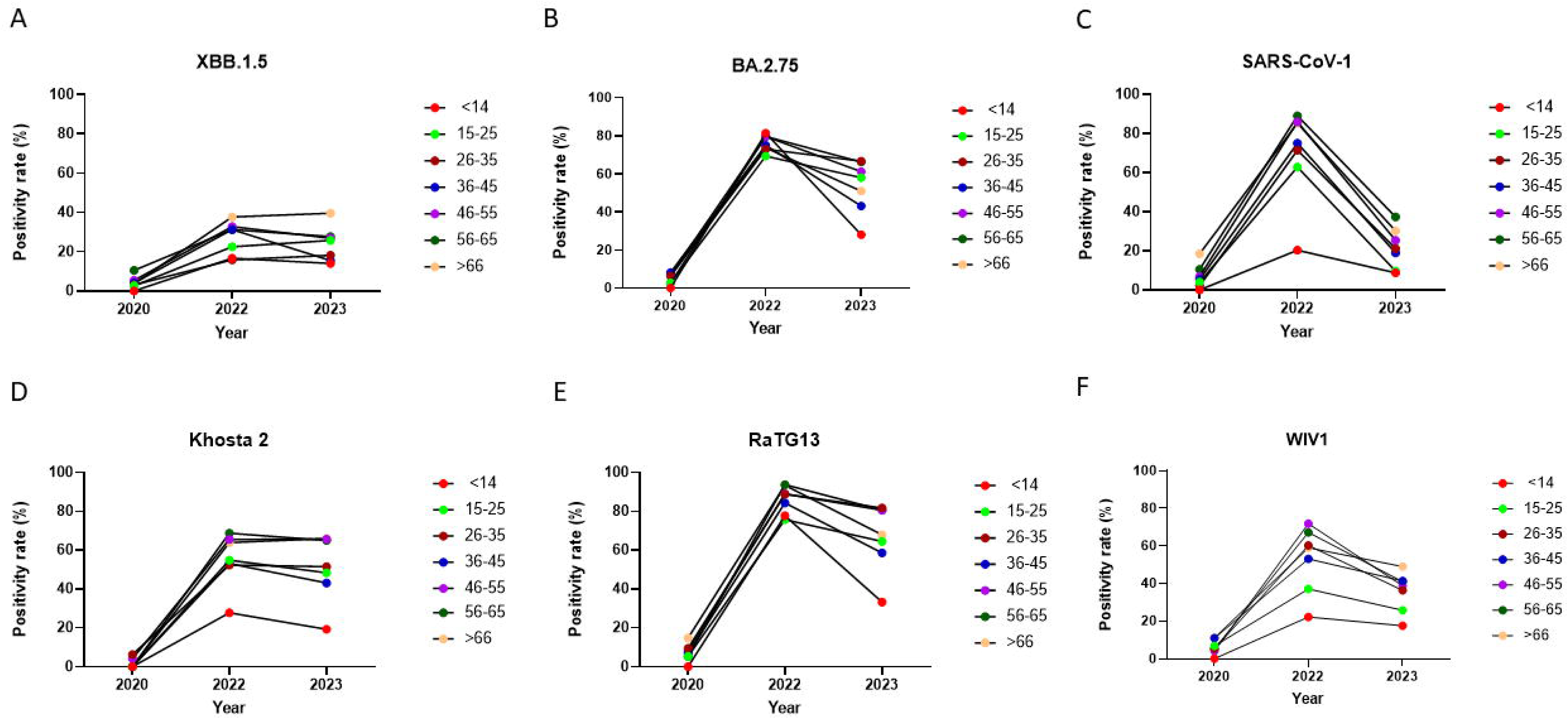
Changes in positivity rates (HAT titres that correspond to a titre of 1:40) for two omicron variants and four sarbecoviruses over time. Antibody titres were measured against the receptor binding domain (RBD) of XBB.1.5, BA.2.75, Khosta-2, WIV1, SARS-CoV-1 and RaTG13 in 2020 (n=381), 2022 (n=432) and 2023 (n=382) using the HAT assay in different age groups in individuals aged 5 to 80 years of age for XBB.1.5 (A), BA.2.75 (B), SARS-CoV-1 (C), Khosta-2 (D), RaTG13 € and WIV1 (F) at different age groups throughout the years.

By 2023, the positivity rates for all these viruses of clade 1A and clade 1B significantly declined except for positivity rates for XBB.1.5 and Khosta-2 (virus assigned to clade 3, in many age groups (Table 1, Figure 2). While positivity rates for SARS-CoV-1 had significantly declined in all age groups, decline in positivity rates for BA.2.75, RaTG13 and WIVI varied in different age groups.

**Table 1:** Age groups that had significantly declined positivity rates from 2022 to 2023 for omicron variants and selected sarbecoviruses.

| Variant | Age groups | Adjusted p value |
| --- | --- | --- |
| BA 2.75 | 36- 45 | 0.002 |
|  | Below 14 | <0.001 |
| W1V1 | 46-55 | 0.002 |
|  | 56-65 | 0.01 |
| Sars COV 2 | 15-25 | <0.001 |
|  | 26-35 | <0.001 |
|  | 36-45 | <0.001 |
|  | 46-55 | <0.001 |
|  | >66 | <0.001 |
| RaTG13 | 36-45 | 0.015 |
|  | >66 | 0.003 |
|  | Below 14 | <0.001 |

The presence of neutralising antibody titres has shown to protect against infection with SARS-CoV-2 and its variants. As a HAT titre of 1: 80 was shown to detect 99% of samples which had neutralizing antibody titres of ≥ 20 (50% inhibitory concentrations, IC_50_) compared to the gold standard microneutralization assay [18], we proceeded to assess the proportion of individuals with a HAT titre of 1:80 for these viruses, to determine the proportion of individuals who were likely to be protected from infection. As seen with positivity rates at HAT titres of 1:40, children <14 years who were unvaccinated had the lowest positivity rates for all tested viruses except BA.2.75 (Figure 3A-F). For BA.2.75 the highest positivity rate of 66.7% for a titre of 1:80 was seen in children <14 years of age. However, these positivity rates significantly declined in 2023 in all age groups for BA.2.75 and the Sarbecoviruses tested except XBB.1.5.

**Figure 3.**
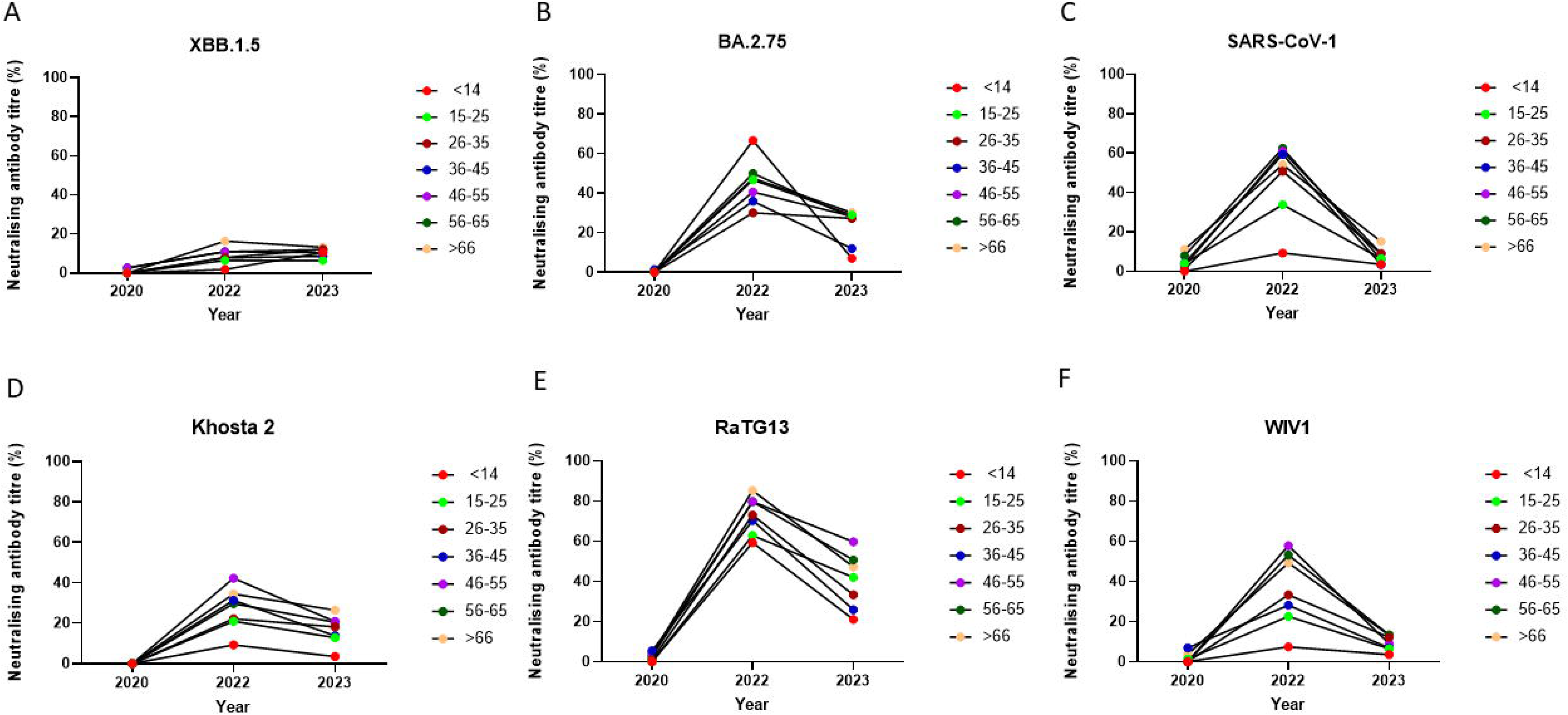
Changes proportion of individuals that have HAT titre of 1:80, that associate with neutralization between 2020, 2022 and 2023 for two omicron variants and four sarbecoviruses over time. Antibody titres were measured against the receptor binding domain (RBD) of XBB.1.5, BA.2.75, Khosta-2, WIV1, SARS-CoV-1 and RaTG13 in 2020 (n=381), 2022 (n=432) and 2023 (n=382) using the HAT assay in different age groups in individuals aged 5 to 80 years of age for XBB.1.5 (A), BA.2.75 (B), SARS-CoV-1 (C), Khosta-2 (D), RaTG13 € and WIV1 (F) at different age groups throughout the years.

### The association between positivity rates and comorbidities

In 2023, we also assessed the presence of comorbidities and the anthropometric measures in the individuals recruited to the study. Therefore, we investigated if the presence of comorbidities such as diabetes, obesity and central obesity would affect positivity rates (HAT titre 1:40) or the presence of antibodies at potentially neutralizing titres (HAT titre 1:80). As children did not have these comorbidities, they were excluded from the analysis, leaving 310 individuals >18 years of age to be included in the analysis. Of the 310 individuals, 17 (5.48%) were classified as underweight as the BMI was ≤ 18.5 kg/m^2^, 101 (32.58%) as healthy, 100 (32.26%) as overweight as their BMI were between 23.9 and 27.5 kg/m^2^ and 92 (29.68%) as obese as their BMI was ≥ 27.5 kg/m^2^. The positivity rates for Khosta-2, WIVI, SARS-CoV-1 and RatG13 were similar in these different groups. However, those who were underweight were significantly more likely (p=0.01) to be positive for XBB.1.5 and BA.2.75 (Figure 4A). Similarly, those with a HAT titre of 1:80, which corresponds to the neutralizing antibody titres, were similar in the different groups for Khosta-2, WIVI, SARS-CoV-1 and RatG13, while underweight individuals, were more likely to have titres of 1:80 compared to other groups (Figure 4B).

**Figure 4.**
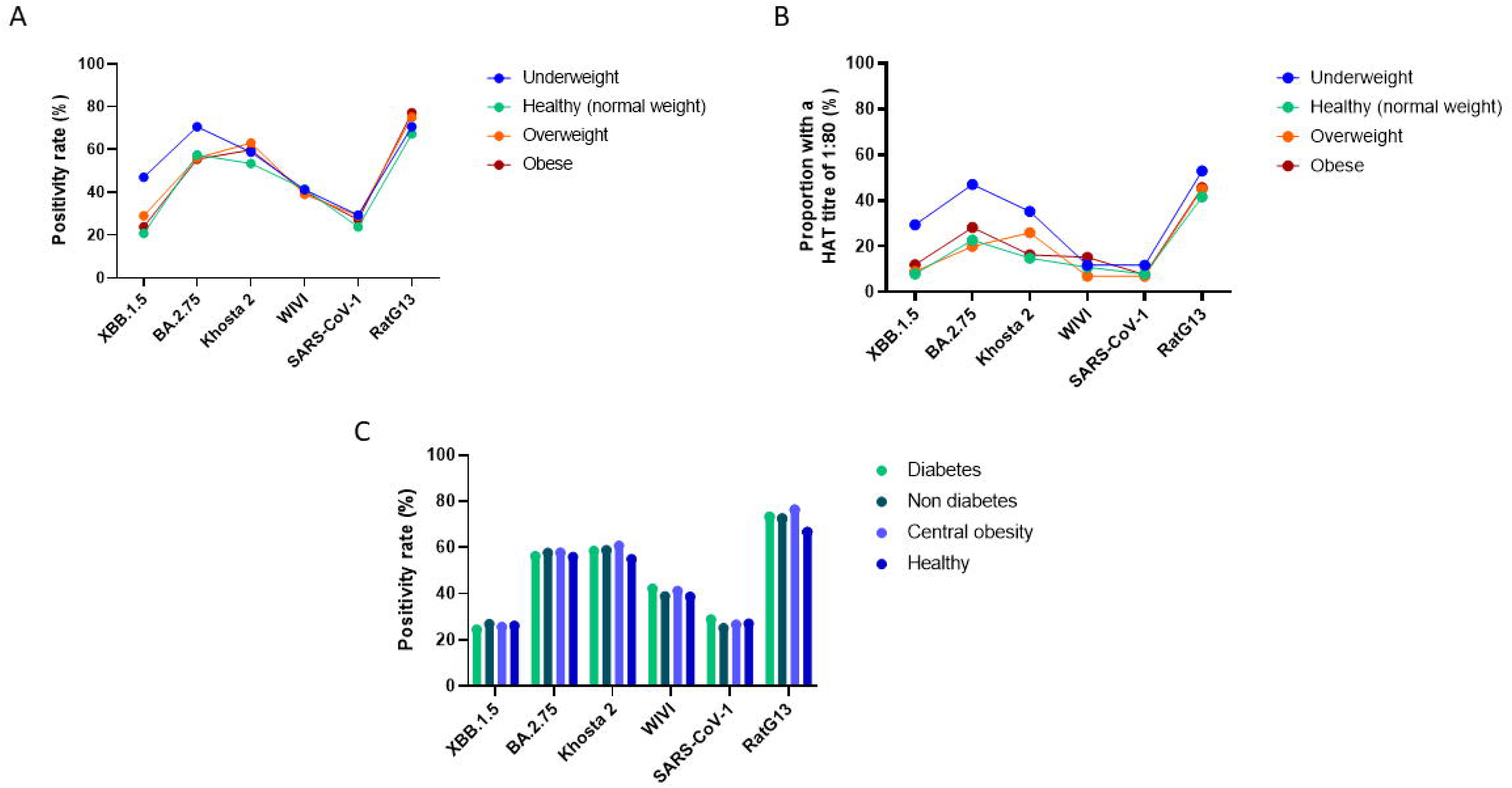
The association between positivity rates for omicron and sarbecoviruses and the presence of comorbidities in individuals. Antibody titres were measured against the receptor binding domain (RBD) of XBB.1.5, BA.2.75, Khosta-2, WIV1, SARS-CoV-1 and RaTG13 In individuals >18 years in 2023 (n=310). The positivity rates (HAT titre of 1:40) (A) and the proportion of individuals with HAT titre of 1:80 for the two omicron variants and the four sarbecoviruses (B) for individuals who were underweight (n=17), healthy (n=101), overweight (n=100) and obese (n=92) are shown. The comparison of the positivity rates for individuals with diabetes (n=135), and without diabetes (n=175), with central obesity (n=199) and with normal waist circumference (n=111) for the different variants are shown in C.

135 (43.55%) individuals had diabetes and 175 (56.45%) were healthy. There was no difference in the positivity rates for any of the tested viruses in these two groups, or between those with central obesity (n=199) and those with normal waist circumference (n=111) (Figure 4C).

## Discussion

In this study, we show that high population immunity rates to SARS-CoV-2 omicron variants and some sarbecoviruses have declined over time, reaching very low positivity levels by September 2023. As expected, the positivity rates for all variants were highest in 2022, following widespread natural infection and vaccination. The highest positivity rates in 2022 and 2023 were seen for RatG13 in all age groups, which is not surprising given that RaTG13 was the most genetically similar virus of those tested [20]. However, the positivity rates for Khosta-2 and WIVI were significantly lower in children <14 years of age, who only had high positivity rates for clade 1A viruses, which included omicron variants and RatG13. This is possibly due to broadly cross-reactive immunity induced in individuals >15 years of age following vaccination and natural infection. In Sri Lanka, while children between aged 12 to 15 received only one dose of a mRNA vaccine (BNT162b2), children <12 years were not vaccinated. An island wide study among children in Sri Lanka, showed that while 99.7% vaccinated and 96.2% had antibodies to SARS-CoV-2, the unvaccinated children had significantly lower positivity rates (67% in unvaccinated vs 95.9% in vaccinated) for ACE2 receptor blocking antibodies, which are associated with protection [21]. However, it was interesting to note that older individuals who were fully vaccinated and were also likely to have been naturally infected, had high positivity rates to both clade 1A and clade 1B sarbecoviruses, whereas children <14 years old only had high positivity rates for clade IA sarbecoviruses. Therefore, a combination of vaccination and infection appears to induce broader immunity to than natural infection alone.

Although positivity rates for all other variants declined from 2022 to 2023, possibly due to lack of exposure during this period, the positivity rates for XBB.1.5 and Khosta-2 remained the same. While infection due to BA.2.75 was seen in the later part of 2022 in Sri Lanka, the dominant variant in 2023 [2], was XBB.1.5 and variants which were descendants of XBB.1.5. Therefore, while a high positivity rate for XBB.1.5 was expected, we were surprised to see similar positivity rates for Khosta-2, which belongs to clade 3 of sarbecoviruses, whereas XBB.1.5 belongs to clade 1A. Therefore, the sequence of infection and vaccination could have an impact on the breadth of the immune response. However, worryingly except for RatG13 where a 60 to 80% positivity rates were observed among adults, positivity rates for other variants were <50% in most age groups.

As neutralising antibodies have shown to protect against severe COVID-19, we also determined the proportion of individuals with titres equivalent to that of 1:80, which an indicator for the presence of antibody titers which can neutralize the tested SARS-CoV-2 variant as previously described [18]. We found that <20% of individuals in all age groups had antibody titres equivalent to 1:80 by 2023. Therefore, the population appears to be susceptible to outbreaks with the emerging variants of SARS-CoV-2 due to low levels of population immunity in 2023. However, we have only assessed antibody responses to different SARS-CoV-2 variants and did not study T cell responses. Broadly cross-reactive T cells have shown to be induced following natural infection and vaccination, which help prevent occurrence of severe disease [22, 23]. In K18-hACE2 transgenic mouse models, it has been shown that mice lacking antibodies to SARS-CoV-2 are protected from infection due to robust T cell responses and functional B cell responses [24]. Therefore, population surveys purely based on assessing antibody responses may not reflect true susceptibility to infection of the population. Therefore, although low population level antibody responses observed by us may indicate some population susceptibility to infection, it does not necessarily indicate the possibility of large outbreaks of severe disease in the Sri Lankan population. However, as T cell responses have shown to wane in the elderly and other susceptible individuals [25, 26], it may be prudent to vaccinate vulnerable individuals with vaccines that protect against the emerging variants [4, 5].

As obesity and diabetes are risk factors for severe COVID-19 [27], we sought to investigate if the positivity rates and antibody titres that associate with neutralizing antibody titres (1:80), varied in individuals with different comorbidities. We found that underweight individuals (BMI ≤ 18.5 kg/m^2^), were significantly more likely to be positive for XBB.1.5 and BA.2.75, which were the variants that circulated in 2022 and 2023. There was no difference for positivity rates for other variants. Although the reasons for these observations are not clear, the number of individuals who were underweight (n=17) was lower compared to other BMI categories, which could have possibly skewed the results.

In conclusion, we observed that population immunity to omicron variants and selected sarbecoviruses had significantly declined from 2022 to 2023 in Colombo, Sri Lanka. Given that <50% of the population had positivity rates for most variants, there is a large population of susceptible individuals in the community. However, T cell responses are known to be important in preventing severe disease, which was not assessed in this study. Therefore, the low antibody positivity rates may not necessarily reflect high susceptibility of the population to severe disease. However, as Sri Lanka did not roll out any booster doses with the latest variants and due to the absence of specific antiviral treatment for SARS-CoV-2, it would be important to consider immunizing vulnerable individuals in the community, especially with those who are likely to have impaired T cell responses, to prevent development of severe disease.

## Statements

### Data availability statement

All data is available within the manuscript, figures and the supporting files.

### Ethics approval statement

Ethics approval for these studies was obtained by the Ethics Review Committee of the University of Sri Jayewardenepura.

### Patient consent statement

All study participants provided informed written consent.

### Permission to reproduce material from other sources

Not applicable.

### Clinical trial registration

Not applicable.

## Acknowledgements

Not applicable.

