## Supplementary tables for "Changes in population immunity to omicron SARS-CoV-2 variants and selected Sarbecoviruses from 2020 to 2023 in urban Colombo, Sri Lanka"

**Supplementary data**

**Table 1.** Number of individuals in each group, in each year

| Age group | Number of individuals’ n (%) | | |
| --- | --- | --- | --- |
|  | 2020 (total n=381) | 2022 (total n=432) | 2023 (total n=382) |
| <14 | 1 (0.26%) | 54 (12.5%) | 57 (14.92%) |
| 15-25 | 74 (19.42%) | 62 (14.35%) | 31 (8.12%) |
| 26-35 | 95 (24.93%) | 63 (14.58%) | 33 (8.64%) |
| 36-45 | 73 (19.16%) | 64 (14.81%) | 58 (15.18%) |
| 46-55 | 73 (19.16%) | 64 (14.81%) | 67 (17.54%) |
| 56-65 | 38 (9.97%) | 64 (14.81%) | 83 (21.73%) |
| >66 | 27 (7.09%) | 61 (14.12%) | 53 (13.87%) |
